# Antimicrobial Prescribing Practices at Dermatology Outpatient Departments in Tertiary Care Hospitals: A multi-centered, Cross Sectional Study

**DOI:** 10.1101/2022.06.19.22276594

**Authors:** Nagina Sultana, Jannatul Ferdoush, Fatema Johora, Sharif Mohammad Towfiq Hossain, Arifa Hossain

## Abstract

**Background:** The purpose of this study is to evaluate antimicrobial prescribing behaviors in the dermatology outpatient department.

**Methods:** This multi-centered, cross-sectional study was conducted at dermatology outpatient department of three tertiary care hospitals in Chittagong division, Bangladesh during the period October, 2021 to April, 2022.

**Results:** During the study period, 463 prescriptions were obtained. Over half of the participants were female (57%) and between the ages of 21 to 40 years. The majority of patients were diagnosed with eczema (19%), acne (17%), and scabies (15%). Most commonly prescribed antimicrobials was Azithromycin (22%), followed by Permethrin (16%) and Erythromycin (7%). In most cases, antimicrobials were recommended for more than two weeks (60%). Nearly half of the antimicrobials (53 %) were recommended as combination of systemic and topical route.

**Conclusion:** Current study found that dermatologists frequently recommended broad-spectrum antimicrobials for extended periods of time, which may lead to antimicrobial resistance. There is a need for continuing medical education on the appropriate use of antimicrobials, which will result in the effective management of skin diseases.

## Introduction

In Bangladesh, prescribing of different types of medicines are generally irrational.[1.2]. Especially, prescribing antimicrobials at different levels and conditions were found inappropriate in different researches[2, 3] Dermatologists frequently prescribe antibiotics in clinical settings for a multitude of purposes. Because antibiotics constitute a vital point of contact between the physician and the patient, there is an increasing need for their prudent usage. Inappropriate use of antimicrobials contributes to the emergence of resistance [4, 5]. Overuse of antimicrobials has led to development of resistant strains, which has become a global problem [6].

Resistance to antibiotics is a rising concern for the efficacy of therapy in dermatologic and infectious diseases [7]. Dermatologists often prescribe antibiotic regimens that last several months [8-10]. Skin problems account for roughly half of all diseases in the developing world, impacting well over 60% of the community [11]. Almost 80% of the total diseases were scabies and pyogenic infections. Children are particularly vulnerable to skin infection [12]. A community-based survey of South-Asian Ameria reported that fungal infections were common (11%) following acne and eczema [13]. The majority of skin ailments in dermatology OPD are cutaneous infections. Bacteria, mainly gram-positive bacteria, cause the bulk of these cutaneous infections. Gram negative bacteria were responsible for some of these infections [14]. The gram-positive bacteria such as streptococcus pyogens, staphylococcus aureus, and propionobacterium acnes caused an overwhelming number of infections. Gram-negative bacteria such as klebsiella, Bartonella sp, p. aeruginosa, rhinoscleromatis, pasturella multocida, Capnocytophaga canimorsus and vibrio vulnicus produced relatively fewer cutaneous infections [14]. Ringworm and oral candidiasis were the two fungal infections with the highest prevalence [15].

Impetigo contagiosa, folliculitis, acne, and erythema are bacterial skin infections where topical therapy is effective. In cases of severe staphylococcal and streptococcal infections, STDs, secondary bacterial infections, acne vulgaris, rosacea, and dermatosis, where bacterial antigens play a pathogenic role, systemic treatment is essential (psoriasis espguttate psoriasis). Clindamycin, ciprofloxacin, metronidazole, fusidic acid, and mupirocin are often applied topically in dermatology OPD [9]. Owing to their strong anti-inflammatory actions, oral antimicrobials are widely recommended for acne, rosacea, and various inflammatory disorder [16, 17].

The World Health Organization has long emphasized the necessity of proper antimicrobial administration and use, which is critical in preventing AMR. Globally, 50% of drugs are incorrectly prescribed, whereas 50% of medicines are misused [18]. As previously stated, irrational antimicrobial prescribing is a universal problem that occurs frequently in clinical practice. Reasons that lead to irrational antimicrobial prescribing include a lack of information about microorganisms, unethical medical marketing, and irrational prescribing behaviors among doctors [19].

This antibiotic use has clinical implications, such as the emergence of resistance [20]. Oral antimicrobial overuse is also linked to major changes in the microbiome, and past antimicrobial exposure is linked to an increased risk of pseudomembranous colitis and other antimicrobial resistant diseases [21,22]. Furthermore, persistent antimicrobial usage has been related to greater risk of colon cancer and breast cancer in some studies [23, 24] which is likely to be mediated by disturbance of the microbiome.

Antibiotic resistance can be controlled by conducting periodic antibiotic-containing prescription audits to analyze and amend. It provided an important educational tool to reduce antimicrobial overuse and misuse, minimize adverse effects, essential drug selection, improve therapeutic efficacy, reduce adverse effects, minimize treatment cost, deliver useful feedback to clinicians, and estimate community drug need, all of which had a positive impact on treatment standards [25]. This, in turn, will improve prescribing habits and improve the efficient management of prevalent skin disorders at a lesser cost.

The purpose of the current study was to describe dermatologists’ prescribing patterns and appropriate use of antimicrobials in order to create baseline data that will assist dermatological prescribers in providing reasonable treatment to their patients.

## Materials and Methods

### Study place and duration

A cross-sectional, multi-centered, descriptive study was undertaken in the outpatient department of Dermatology in three govt. affiliated three tertiary care hospitals (BGC Trust medical College, Marine City Medical College and Chattogram Maa-O-Shishu Hospital Medical College) in Chittagong division, Bangladesh from October 2021 to May 2022. Ethical approval was taken from Institutional Review Board (IRB) of BGC Trust Medical College. Study procedure: Total of 463 prescriptions containing antibiotics were recruited in the study The study included new patients who came to the dermatology outpatient department, but not those who were admitted during their visit. The prescriptions were thoroughly reviewed, and relevant data was collected, including demographic details, diagnosis, and the name, duration and formulation of the antimicrobials prescribed for the infection. All of the information was gathered, collected, tabulated, and assessed. Data quality was ensured through extensive checks for duplicate and missing values, as well as prompt correction of any errors. The information was presented as a percentage and also as a total figure. A Microsoft Excel 2010 spreadsheet was used to analyze the results.

## Results

The study comprised 463 prescriptions from dermatology outpatient department patients that included antimicrobials. Demographic characteristics of study population are illustrated in Table I. The majority of the patients (58%, 269/463) were between the ages of 21 to 24 years. Among them 57% (266/463) were female patient.

**Table I:** Participants’ demographic information

| Demographic characteristics | No of patients (n=463) | Percentage (%) |
| --- | --- | --- |
| Age (years) |  |  |
| <20 | 58 | 13 |
| 21-40 | 269 | 58 |
| 41-60 | 87 | 19 |
| >60 | 49 | 11 |
| **Gender** |  |  |
| Male | 197 | 43 |
| Female | 266 | 57 |

Table II showed that most common indication of antimicrobial prescribing were eczema 19% (87/463), acne vulgaris 17% (79/463), scabies 15% (76/463), allergic Contact dermatitis 8% (37/463)

**Table II:** Dermatological conditions indicated for antimicrobials (n=463)

| Dermatological Indication | Total No of patient |  |
| --- | --- | --- |
|  | Frequency | Percentage (%) |
| Eczema | 87 | 19 |
| Acne vulgaris | 79 | 17 |
| Scabies | 76 | 15 |
| Allergic Contact dermatitis | 37 | 8 |
| Impetigo | 16 | 3 |
| Insect bite | 5 | 1 |
| Folliculitis | 9 | 2 |
| Atopic dermatitis | 19 | 4 |
| Tinia corporis | 24 | 5 |
| Tinia cruris | 18 | 4 |
| Pityriasis versicolor | 17 | 4 |
| Prurigo Simplex | 7 | 2 |
| Seborrheic dermatitis | 23 | 5 |
| Furuncle and carbuncle | 4 | 1 |
| Psoriasis | 18 | 4 |
| Vulvovaginal candidiasis | 11 | 2 |
| Alopecia areata | 2 | .4 |
| Candida intertrigo | 3 | .6 |
| Paronychia | 8 | 2 |

Table III showed that most frequently used antimicrobials were Azythromycin 22% (104/463), permethrin 16% (76/463), Erythromycin 7% (33/463).

**Table III:** Antimicrobials prescribed in dermatology out-patient department (n=463)

| **Antimicrobials** | **Frequency** | **Percentage (%)** |
| --- | --- | --- |
| Azythromycin | 104 | 22 |
| Permethrin | 76 | 16 |
| Erythromycin | 33 | 7 |
| Doxycycline | 23 | 5 |
| Cefuroxime+Clavulanic acid | 6 | 1 |
| Gentamicin | 5 | 1 |
| Flucloxacilin | 21 | 5 |
| Mupirocin | 25 | 5 |
| Clotrimazole | 4 | 1 |
| Acyclovir | 11 | 2 |
| Metronidazole | 11 | 2 |
| Voriconazole | 16 | 3 |
| Luliconazole | 15 | 3 |
| Fluconazole | 19 | 4 |
| Ketoconazole | 23 | 5 |
| Itraconazole | 12 | 3 |
| Terbinafine | 13 | 3 |
| Clindamycin | 14 | 3 |
| Tolnaftate | 6 | 1 |
| Cefuroxime | 11 | 2 |

Table IV showed the distribution of antibimicrobials according to the routes of delivery. A combined route (systemic and topical) was given in 53% (232/463) encounters, followed by an oral route in 25% of cases. In terms of duration of usage, 60% (277/463) of participants prescribed antimicrobials for less than one week, whereas 40% (186/463) for more than two weeks.

**Table IV:** Routes of administration of antimicrobials (n=463)

| Routes of administration | Frequency | Percentage |
| --- | --- | --- |
| Oral | 118 | 25 |
| Per-enteral | 00 | 0 |
| Topical | 103 | 22 |
| Combination (systemic and topical) | 232 | 53 |
| Duration of use |  |  |
| >2 weeks | 186 | 40 |
| ≤1 weeks | 277 | 60 |

## Discussion

In light of the widespread global antimicrobial resistance, irrational antibiotic prescribing is a major source of concern. The foremost goal of dermatological treatment is to prescribe the least and safest antimicrobials possible. The problem is exacerbated by the illogical and inappropriate usage of antimicrobials. Therefore, the objective of the study was to evaluate prescribing pattern of antimicrobials by dermatologist. In this study, a high number of patients (58%) were between the ages of 21 and 24, and more than half of the participants (57%) were female.

In the current study, eczema accounted for 19% of antimicrobial prescriptions, followed by acne vulgaris (17%), and scabies (15%). Bahelah SO et al study also revealed similar to this study [26] According to Jannat et al study, scabies was the most frequent reason for dermatological disease seen in hospitals’ outpatient departments, followed by dermatitis, folliculistis, urticaria, impetigo, psoriasis, acne, and melasma. [27]

The most frequently antimicrobials recommended in this study were azythromycin (22%), permethrin (16%), and erythromycin (7%). The most regularly recommended antibiotics were Macrolids (33.2 %), followed by penicillin (24.7%), and according to a study conducted by Bahelah SO et al.[26] Macrolides are frequently recommended, maybe because they have fewer adverse effects. The most commonly recommended antimicrobial medicines for skin infections are beta-lactam medications (penicillin, cephalosporin) and Macrolides, according to prior studies [28-30].

In this study, dermatologists preferred the combination route (systemic and local) 53 %, followed by oral route, which was recommended in 25% encounnters. Concomitant usage systemic and topical antimicrobials for acne should be avoided, according to several studies, because it increases the risk of multidrug-resistant organisms and gives no further benefit. [31,32]

Topical antibiotics provide a number of advantages, including a reduction in oral antibiotic misuse, a reduction in bacterial resistance, and the avoidance of systemic toxicity and side effects. [33] Furthermore, several study suggest that topical antimicrobials are more efficacious than systemic antimicrobials of skin infections like impetigo, and that they should be used first.[konnoing] [34] The use of topical antimicrobial is limited in the current study, which is regarded unreasonable, yet in our circumstance, many patients presented with widespread lesions as well as poor hygienic environment.

As a result, dermatologists have no choice except to prescribe oral antimicrobials, either alone or in combination with topical antimicrobials, in these circumstances. Prescribers should stop prescribing fixed dose combinations of topical medicines containing antimicrobials, antifungals, and steroids, which could minimize antimicrobial use in dermatology.

In terms of antimicrobial use duration, the current study found that 60% of antimicrobials were prescribed for more than one week. In contrast to the current study, the Bahelah et al [26] finding revealed the duration of antimicrobial use, with the majority of cases being treated for one week (78.4 %).

Antibiotic use for an extended period of time is one of the variables that leads to the emergence of resistant bacteria. This finding is comparable to the findings of Anuj Kumar Pathak et al study [35].

The study has a few shortcomings. Only antimicrobial prescribing in dermatological outpatients was observed. Dermatology patients admitted as in-patient department were not included. Safety of antimicrobial use was not investigated.

## Conclusion

Dermatologists are increasingly prescribing broad-spectrum and long-acting antimicrobials, according to the findings of current study, placing patients at risk for adverse effects and perhaps leading to antimicrobial resistance. Dynamic educational approaches should be used in order to avoid needless and prolonged antimicrobial use. This findings will help dermatologists to prescribe with more precision. In order to determine the effectiveness of this approach in terms of patient outcomes and antibiotic stewardship, further research is required.

## Data Availability

yes

